# A dynamic model for Covid-19 in Brazil

**DOI:** 10.1101/2020.05.10.20097550

**Authors:** Rubens Lichtenthäler Filho, Daniel Gomes Lichtenthäler

## Abstract

A dynamic model for the current coronavirus outbreak is presented. The most important parameters are identified which determine the number of cases progression. Results of a numerical simulation are compared with existing data of the number of COVID-19 cases and deaths in Sao Paulo and Brazil. On the basis of these results we display the effect of social distancing measures taken so far, which flattened the infection curve. A simple three steps procedure is proposed to predict changes in the evolution of the epidemics and we discuss the importance of serological representative surveys to relate the epidemic time to the real time. A criteria to start relaxing social distance measures is suggested.

## 2 Introduction

Epidemiological models are an important tool to predict the time evolution of outbreaks and to guide official control measures. There is a common belief among physicians and professionals of the field that the reliability of model predictions is extremely limited by the high degree of sub-notification usually present in epidemiological data. Indeed, the number of individuals infected by SARS-CoV-2 seems to be largely underestimated by official reports all over the world [1]. Nevertheless, comparison between model predictions and epidemiological data is the only way to determine the parameters involved in the calculations. The extent to which these parameters are affected by sub-notification factors is a crucial point to assess the model predicting power.

The multiplication process of the number of cases in an epidemics is primarily determined by the infection rate or reproduction number (R_0_). This dimensionless quantity is defined as the average number of new infections caused by each infected person. In the case of SARS-COV2 outbreak in Sao Paulo this number is estimated to be between 2-6, if no social distancing measures are taken and the epidemics is left to follow its natural course. The infection rate (R_0_) is the parameter that controls the number of new infections in a certain moment given the number of infected people in the days before. The assumption that in the early phases of the outbreak the new number of cases is completely determined by the product between Ro and the current number of infections implies directly that the epidemics curve will be an exponential whenever R_0_ > 1. As R_0_ increases, the steepness of the exponential also increases. The infection rate parameter R_0_ can be reduced by adopting measures of social distancing and quarantine [1, 2]. A decrease in the social interaction rate and hygiene measures have the potential to strongly decrease Ro reducing the steepness of the exponential curve and flattening the infection curve. For *R*_0_ ≤ 1 the infection curve becomes flat and the epidemics is controlled.

Another important parameter in the dynamics of the epidemics is the fraction of the population immune to the virus (as there is no vaccine, we assumed this to be the healed fraction of the population). Its complementary quantity is fraction of the population susceptible to the virus (T). T is given as the ratio between the number of susceptible people and the total population and decreases in time as more and more people are being healed reducing the virus transmission probability. The fraction of susceptible people should decrease in time, assuming that all the healed population became immune although recent findings indicate that the latter assumption may not be rigorously valid [3].

Other important parameters are the incubation time (we use it as a reasonable proxy for the peak of the probability of transmission once infected by the virus [4]) and the healing time. These parameters basically determine all the dynamics of the epidemics.

In the next sections we will present details of the model and compare the results of the numerical simulation with epidemiological data.

## 3 Numerical Model and results

The algorithm calculates the new number of infections in a certain day, given the number of individuals infected in the previous days. The total number of individuals infected in a day n is obtained by the equation below:

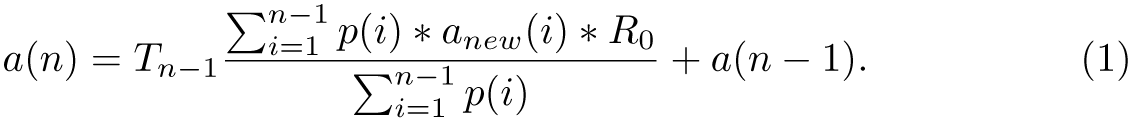

Where *a_new_*(*i*) stands for the new infections in each day *i*. Note that the number of new cases in day n, given by the first term in the right side of Eq.1, is obtained from the contribution of all previous new cases since the beginning of the epidemics (day 1) up to day n-1, weighted by a probability *p*(*i*) which is a function of time. The probability *p*(*i*) was taken as a Gaussian distribution centered in a distance backwards from day n equal to the incubation time (*τ_inc_*) with a given width (*w*). Every infected person will contribute to a certain number of new infections with a probability that depends on how far backwards it is from day n. This probability increases from day 1 up to a maximum in the incubation time day, decreasing down to day n-1. The Gaussian is normalized to 1 by the factor in the denominator.

The factor (*T_n_*) multiplies the summation in the right side of Eq.1 and is an important one. It is defined as 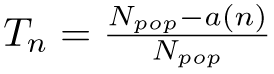, where *N_pop_* is an arbitrary total population. As *a*(*n*) is a function of time, *T_n_* will also be a function of time starting from *T* =1 (*a*(*n*) = 0) dropping down as the total number of infections *a*(*n*) increases.

The number of new cases in day n has to be added to the total number of cases until day n-1 to obtain the cumulative number of infections.

In Figure 1 we plot the cumulative number of individuals infected given by Eq.1 versus the time measured in days, starting in the day of the first reported infection in Sao Paulo (also the first reported case in Brazil) Feb 26, 2020. We present the results of the simulation for three different infection rate parameters (solid lines) *R*_0_ = 3.5, 1.9, 1.4 compared to the reported number of infections in Brazil (blue circles). The other parameters on these calculation are: incubation time *τ_inc_* =4.4 days [1], width of Gaussian distribution *w* =2.2 days and a total population of 200 million people. The reported number of cases in Brazil (blue circles) have been multiplied by a constant factor 7.14 which stems from the estimated undocumented factor of 86% as reported in [1]. With this correction factor our mortality rate is around 1%.

**Figure 1:**
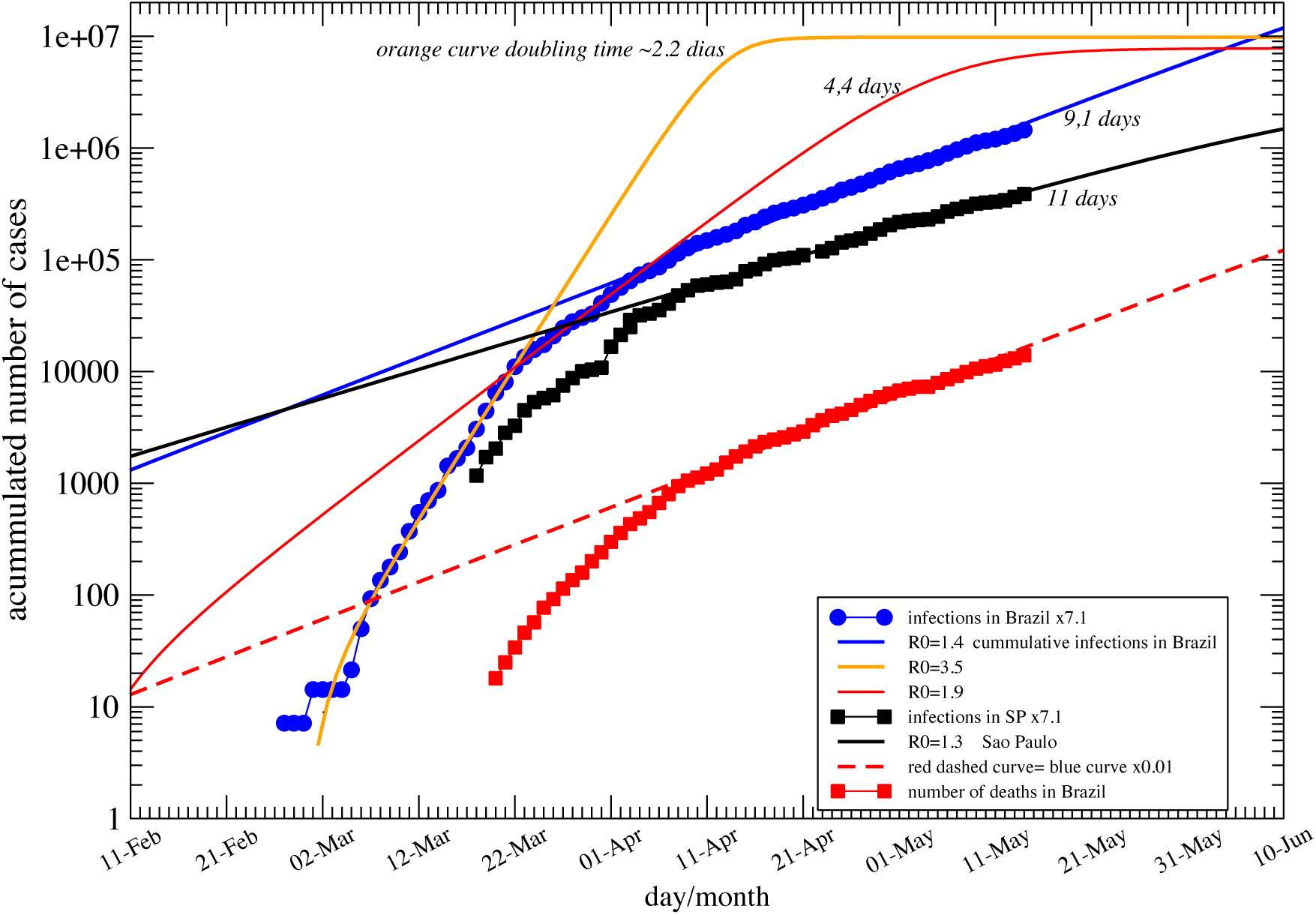
Contamination curves (solid lines). Number of cases in Brasil (blue circles). Number of cases in Sao Paulo (black squares), number of deaths in Brazil (red squares).

The black squares are the Sao Paulo state data compared to the black curve for *R*_0_ = 1.3.

The curves are plotted in logarithm scale. This is an important remark since, by taking the log of an exponential, any terms multiplying the exponential become constant and are washed out when the derivative is taken. As *R*_0_ is related to the derivative (slope) of the curves, this means that the determination of *R*_0_ is independent of constant multiplicative factors in the exponential. As a consequence, even if the data are underestimated as they are, that would not affect *R*_0_, as long as the underestimating factor stays constant in time.

In Figure 1 we clearly see 3 regimes in the time evolution. From Feb 26 until around March 23 data are following the orange curve corresponding to *R*_0_ = 3.5. From 23 of March up to 08 of April it follows *R*_0_ = 1.9 (red curve) and after that until May 12 it follows *R* _0_ = 1.4. The doubling time is indicated in the figure for the orange, red, blue and black curves. The relation between *R*_0_ and the time to double the number of cases (doubling time *τ_d_*) in the ascending part of the curve can be estimated as:

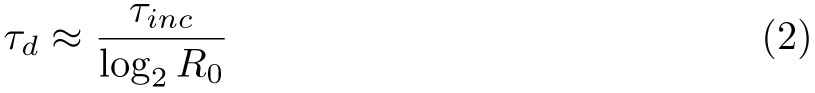

where *τ_inc_* is the incubation time.

The correspondence between the epidemic time and the real time is not trivial. This relation is of great interest since it would allow us to predict how far we are from the peak (and also from the end) of this outbreak. Random surveys performed in representative samples of the population could determine the percentage of the population which has already been in contact with the virus, providing and estimation the *T_n_* factor which, by its turn, would give an idea of our real position in the epidemic time. Unfortunately such surveys are not available in Brazil yet.

In Figure 2 we compare the number of new cases reported every day in Brazil (data goes up till May 12) to the simulation results for *R*_0_ = 1.4 and the same parameters as quoted in Figure 1 (blue curve). All curves have been calculated with the same *R*_0_ parameter but normalized by different constants and shifted in time by different amounts. This corresponds to assuming different total population in the sample. We see that all curves are consistent with the data but give different predictions for the position of the peak indicating that it is not possible to predict exactly when the maximum of infection will be attained [5]. Nevertheless, the percentage of the population infected at the peak of the outbreak is completely determined by *R*_0_ (as well as the percentage of population contaminated at the end of the outbreak), this is shown in Figure 3.

**Figure 2:**
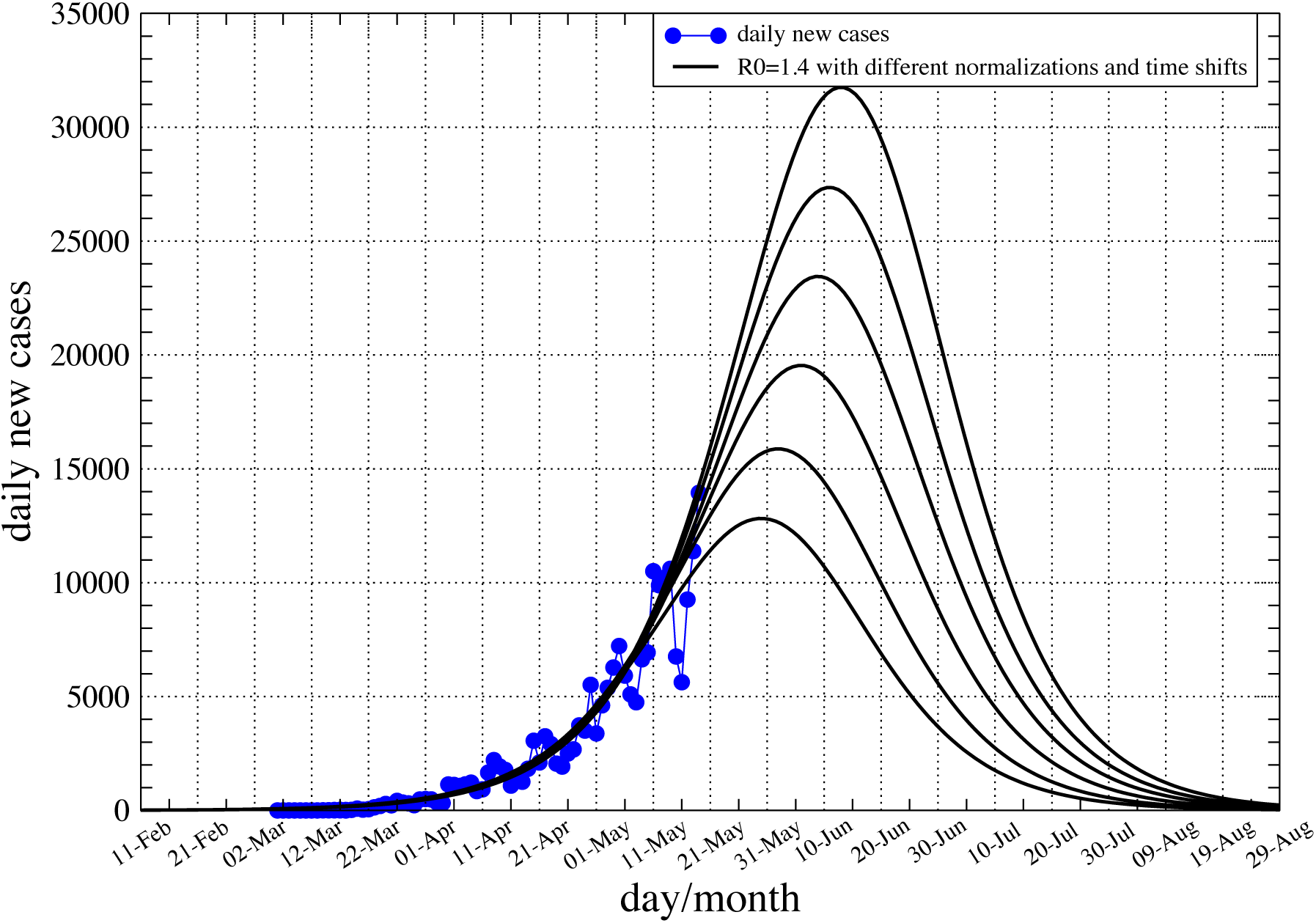
Reported daily number of new cases in Brazil (blue dots), solid line is the result of the simulation with the same parameters as in Figure 1 (blue curve) with different normalizations and time shift factors.

Another important factor that has to be taken into account is the delay in the reporting time. There are indications that, in Brazil, the reported data can be delayed by up to 14 days implying that the real position in the curve of Figure 2 should be shifted to the left by this amount of time.

In Figure 4 we show the results of two calculations. The red curve is the cumulative number of cases and the black curve is the daily infection curve for *R*_0_ = 1.4 and population of 10 million. For *R*_0_ = 1.4, 51% of the population is contaminated [6]. The vertical dashed line shows the position of the peak and its correspondence on the total number of cases curve. We see that at the peak of the outbreak about 3.3 million cases (red curve) in a total of 5.1 million are reported. It means that, in the peak of the new infection curve, about 65% of the final number of cases is reached, corresponding to 33% of the total population. The latter corresponds to the herd immunity factor (*R*_0_ × *T_n_* ≤ 1) as quoted in [6]. This point will be discussed in more details in the next subsection. At this point the number of new infections starts to decrease. The total number of cases still continues to increase and reaches about 88% of the total number of cases (5.1*millions*), approximately 14 days after the peak. If we consider the median healing time of 14 days, this corresponds to the peak of the healed daily cases curve. We propose this as a criteria to start relaxing quarantine and social distance measures.

**Figure 4:**
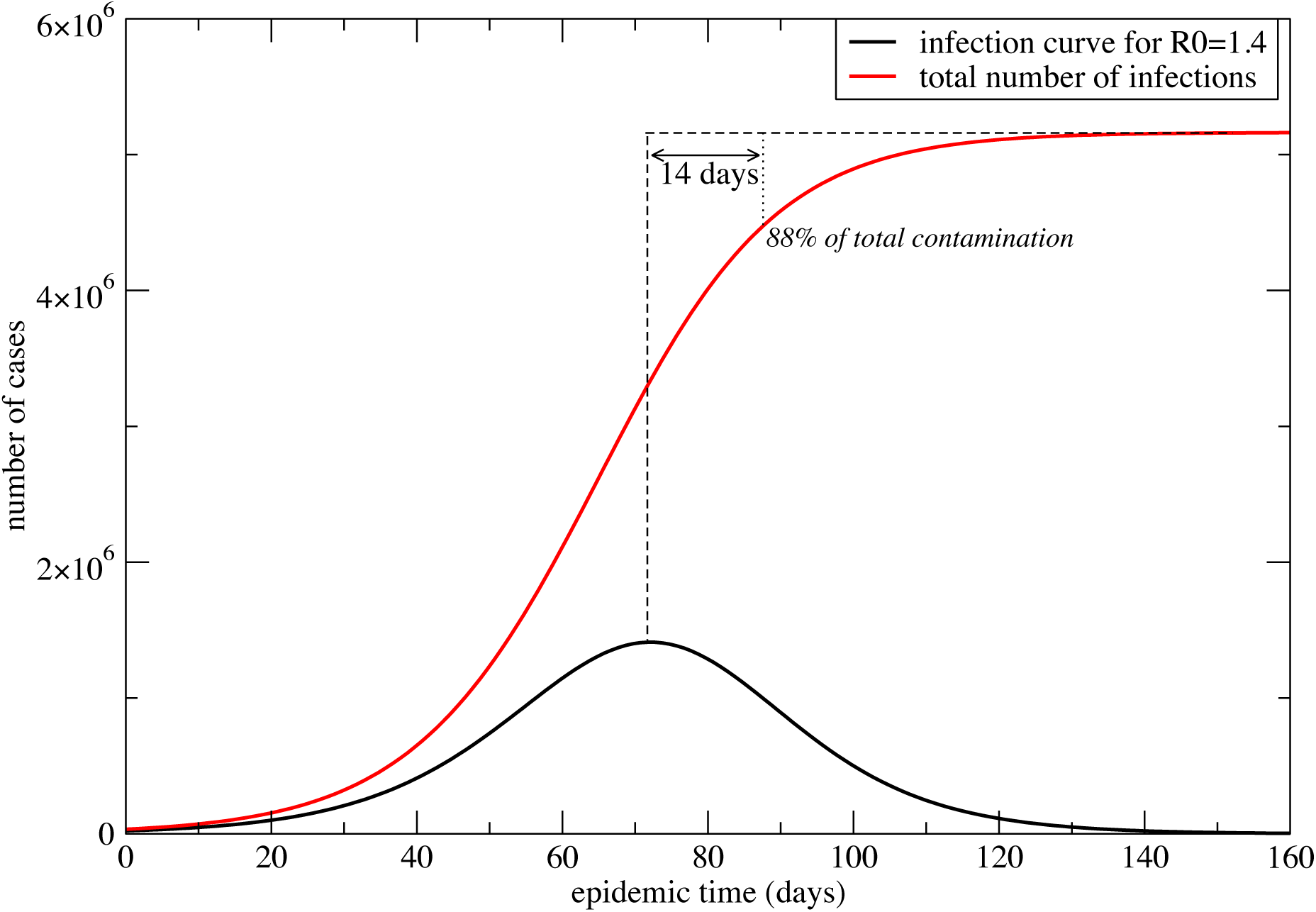
Integrated number of infections (red line) and total number x time. Calculation performed for *R*_0_ = 1.4 and 10 million population.

### 3.1 Herd immunity

Herd immunity can be understood as the percentage of the population that needs to be immune in order to slow down the spread of the virus. *T_n_* is the fraction of the population susceptible to infection and is a decreasing function of time (as discussed above). Then, for a given *R*_0_, as *T_n_* will reach a value below which 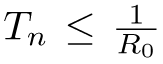 and, as the effective infection rate (*R*_0_ × *T_n_*) turns out to be smaller than one, the epidemic becomes controlled. We can define 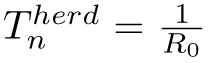 and see that herd immunity (1 − *T^herd^*) is determined by *R*_0_ as show in Figure 3. For *R*_0_ = 3.5 one gets 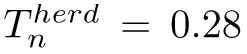, implying that 1 − *T^herd^* = 72% of population is immune. For *R*_0_ = 1.4 the fraction of immunity drops down to 28%. For *R*_0_ = 2, *T^herd^* = 50%. Herd immunity is an important parameter since it gives an idea of the magnitude of the immune population sufficient to stop the exponential epidemic spread (see Figure 3).

**Figure 3:**
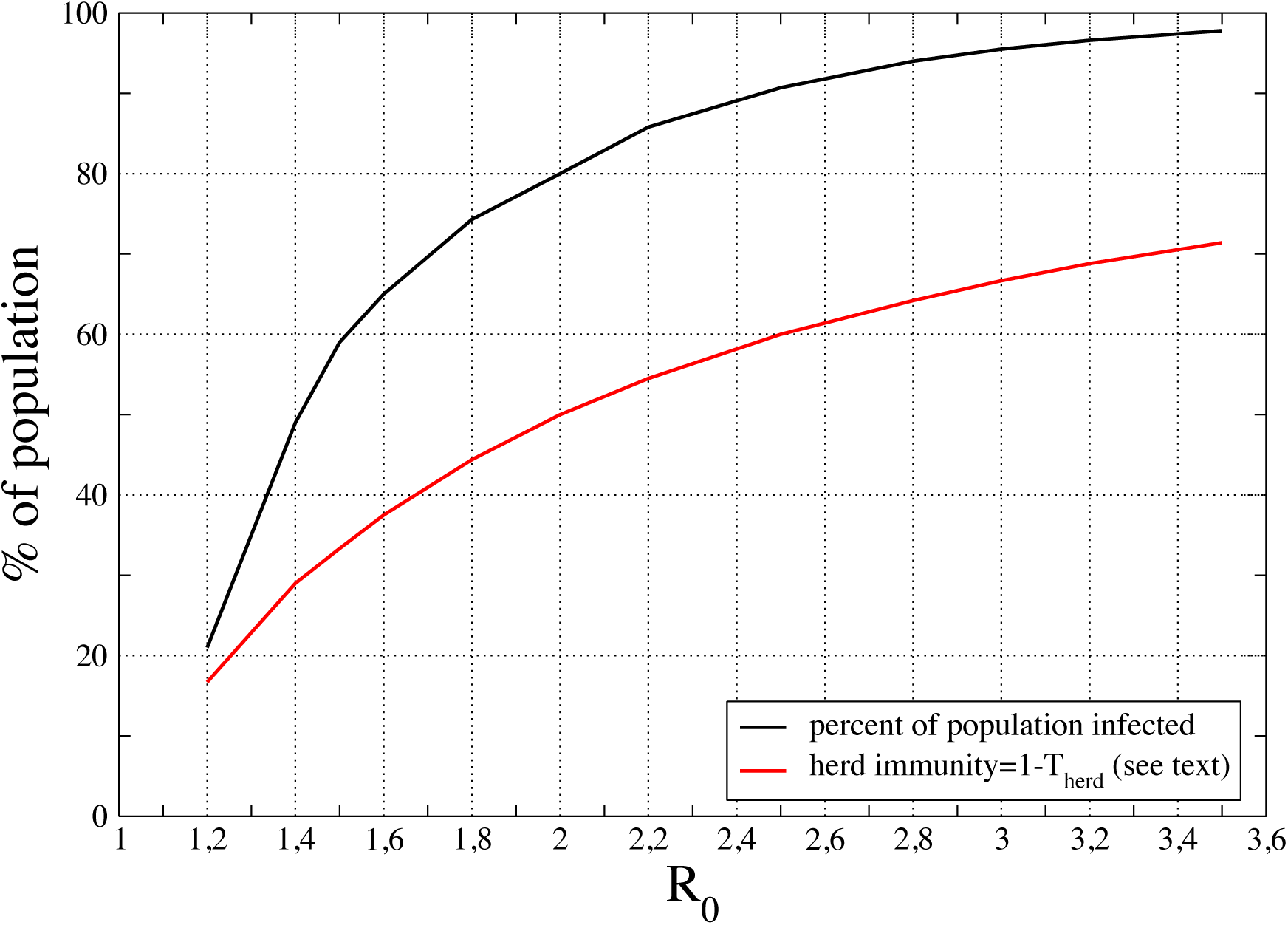
Percent of population contamined and herd immunity as a function of *R*_0_.

Assuming that present Brazil‘s *R*_0_ ranges between 1.4 − 2.0, one would expected 28 − 50% required immunity to stop epidemics, as long as *R*_0_ stays constant in this range.

It seems to be very important to perform random surveys of serological tests in the population to probe the evolution of the fraction of the immune people, giving an idea of how far we are from the herd immunity condition for a given *R*_0_.

On the light of these findings we propose a simple three steps procedure to estimate the herd immunity directly from epidemiological data:

- Determine the infection doubling time directly from epidemiological data.
- Apply the simple formula^1^ that relates *R*_0_ with the infection doubling time to obtain 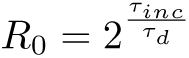 where *τ_inc_* and *τ_d_* are incubation and doubling time respectively.
- Once *R*_0_ is known, the herd immunity can be easily obtained as 1 − 1/*R*_0_.

## 4 Conclusions

We develop a model and a computer code to calculate the dynamics of Covid-19. The model is a simple one but contains all the important ingredients, to be known, the infection rate parameter *R*_0_, incubation time, healing time and the immunity factor (T).

Calculations have been performed for different *R*_0_ factors and comparison with epidemiological data shows that it is possible to determine quite precisely the value of the infection rate *R*_0_ even if the data are sub-notified as long as the sub-notification factor is constant. Comparison of our calculations with data show 3 moments in this epidemics in Brazil so far, with different *R*_0_ factors *R*_0_ = 3.5; 1.9 and 1.4. The observed reduction of *R*_0_ was probably induced by the quarantine and social distancing measures and eventually lock-down adopted by State governments. In Sao Paulo the infection curve is following a little smaller *R*_0_ = 1.3 value. It became clear that the social distancing politics is essential to reduce the total number of cases and to control the epidemics. The total number of infections in the end of the outbreak drops from 97% down to 51% as the infection rate falls from *R*_0_ = 3.5 to for *R*_0_ = 1.4. We show that, at the peak of the contamination curve, about 64% of the total contamination has been reached and 14 days after the peak about 88% of the total contamination took place suggesting the peak of the healing curve as a possible criteria to start relaxing social distance measures. It is important to notice that relaxing these measures will increase *R*_0_ in an unpredictable way, so relaxation measures should be gradual and followed by continued monitoring the epidemic curves so that subsequent relaxing or restrictive measures can be taken as needed.

Conducting random surveys to determine the percentage of infected people in the population is a critical measure to determine the time position in the epidemic curve, which would by its turn, provide an estimation of the distance to the peak. The fraction of infected people (1-*T_n_*) in the population is an important parameter to estimate how far we are from the herd immunity condition (for a given *R*_0_) and to guide the implementation of relaxation in the social distancing measures. This percentage can be obtained from serological tests performed in random representative samples of the population. Otherwise, if there is no control of the percentage of infected people, it turns out very difficult to make reliable predictions of how far we are from controlling the epidemics.

In order to implement these procedures we have to keep in mind that all the official reports provide a delayed picture of the epidemics, the delay time depending on several factors such as incubation time, time to hospitalization, reporting time, etc. An evaluation of the reporting delay time is also highly desirable in order to provide a reliable estimation of the time position in the epidemic curve.

More radical social distancing measures such as lock-downs (aiming *R*_0_ < 1) should be adopted, based on the analysis of local epidemic data of each city or community (and its health-care capacity) rather than on data from the whole country/state. Epidemiological data taken over the whole country/state are rather inclusive and resulting from the contributions of several smaller out-brakes taking place in different locations at different times. Permanent assessing of the out-brakes evolution in each city is a highly desirable measure to take the situation under control and eventually implement local lock-downs restricted to certain cities for limited periods of time.

## Data Availability

I used available official data from brazilian govern

## Footnotes

1. this formula is the inverse of equation 2.

## Notes

### Competing Interest Statement

The authors have declared no competing interest.

### Funding Statement

University of Sao Paulo
CNPq

